# Magnitude of Undernutrition and Associated Factors among Pregnant Women Attending Public Health Facilities in Goba District, Bale Zone, Oromia, Ethiopia: A Cross-sectional Study

**DOI:** 10.64898/2026.06.05.26354999

**Authors:** Seifu Mohammed Ibrahim, Meron Seyoum Lakew, Abebe Feyissa Amhare, Dursa Hussein, Habib Kedir, Haris Abdulbesit

**Affiliations:** Department of Public Health, Ginnir General Hospital, Oromia Regional Health Bureau, Ginnir, Ethiopia; Department of Public Health, Goba District Health Office, Oromia Regional Health Bureau, Goba, Ethiopia; Department of Public Health, College of Health Science, Salale University, Fitche, Ethiopia; Department of Nursing, Kuyu General Hospital, Oromia Regional Health Bureau, Gerba Guracha, Ethiopia; Department of Public Health, International Rescue Committee Adama Field Office, Adama, Ethiopia

**Author notes:** Corresponding Author: Seifu Mohammed Ibrahim (SMI).

**Keywords:** Undernutrition, Pregnancy, public health facilities, cross-sectional study, Goba district

## Abstract

**Objective:** This study aimed to assess the magnitude of undernutrition and associated factors among pregnant women attending public health facilities in the Goba district, Bale zone, Oromia Region, Ethiopia, 2022.

**Design:** Institution-based, cross-sectional study design was used.

**Setting:** The study was conducted in selected public health facilities from May to June 2022.

**Participants:** The study population consisted of pregnant women who lived for at least 6 months in the study area and who attended antenatal care follow-up at selected public health facilities during the study period. Pregnant women who lived for less than six months in the study area and those who were critically ill were excluded from the study.

**Results:** 487 respondents participated in this study with a 100% response rate. More than half (50.7%) of pregnant mothers were undernourished. The significant factors associated with maternal undernutrition during pregnancy in this study were mothers with no formal education (AOR = 5.050; 95% CI: 1.470-17.346), a history of illness during pregnancy (AOR = 2.089; 95% CI: 1.246-3.504), and eating frequency of meals less than or equal to three times per day (AOR = 3.292; 95% CI: 1.040-10.42). Poor nutritional knowledge (AOR = 5.588; 95% CI: 2.921-10.689), poor household (HH) wealth status (AOR = 4.774; 95% CI: 2.216-10.285), and mothers who had >= 4 pregnancies were included (AOR = 0.852; 95% CI: 342-0.989).

**Conclusion:** The magnitude of Undernutrition among pregnant women was 50.7%. Significant associations with Undernutrition were found in mothers with no formal education, poor dietary knowledge, a meal frequency of three or fewer times per day, a history of illness during pregnancy, lower and medium household wealth status, and those who had experienced four or more pregnancies while attending antenatal care (ANC) services at public health facilities.

**Strengths and limitations of the study:**

- Comprehensive data collection covers socio-demographic, economic, dietary, and health factors.
- Standardized measurement techniques and adapted tools ensure data reliability and validity
- As a cross-sectional study, a cause-and-effect relationship cannot be established to identify actual predictors
- Reliance on self-reporting for dietary habits and illness history may introduce bias.
- The study excluded pregnant women who were not utilizing ANC services.

## INTRODUCTION

Maternal malnutrition can be defined as a lack of sufficient food or deficiency of a specific nutrient[1]. The immediate causes of maternal malnutrition include inadequate food intake, poor nutritional quality of diet, frequent infections, and short inter pregnancy intervals [2]. Multiple nutrient deficiencies are generally observed in women with low socioeconomic status [3].

A mother’s nutritional status prior to conception, (preconception 3 months before), and immediately afterwards is critical. The fetus is most vulnerable to nutritional deficiencies in the first trimester of pregnancy, often before a woman realizes that she is pregnant [4]. Nutrient needs typically increase more during pregnancy than during any other stage in a woman’s adult life. Additional nutrients are required during gestation for development of the fetus as well as for growth of maternal tissues that support fetal development [5].

Maternal nutritional status and nutrient intake before and during pregnancy can have a profound effect on the health of both mother and child. A varied, nutrient-dense diet is important for both the mother and baby during pregnancy [6]. Nutritional deficiencies are still common problems during pregnancy. Chronic energy deficiency is the result of prolonged food deprivation manifested as low body weight and energy stores [8].

Deficiencies in micronutrients such as thiamine, niacin, copper, selenium, and zinc have supportive influences on pregnancy and affect maternal conceiving capacity by controlling gametogenesis, fertilization, and development of the embryo before implantation [9]. The prevalence of undernutrition in mothers is 462 million worldwide[10], and it has become the most important risk factor for morbidity and mortality and affects hundreds of millions of pregnant women and young children[11]. Improving maternal nutritional status before conception and during pregnancy is essential for improving the birth weight of newborns [3].

Adequate nutrition is essential for women throughout their life cycle, and pregnant women need a variety of diets and increased nutrient intake to cope with extra nutritional requirements during pregnancy and lactation. The use of dietary supplements and fortified foods should be encouraged for pregnant mothers to ensure an adequate supply of nutrients for mothers and fetuses [12]. Therefore, this study was conducted to assess the magnitude of undernutrition and associated factors among pregnant women who attended antenatal care (ANC) follow-up at selected public health facilities in the Goba district, Bale zone, Oromia, Ethiopia.

## METHODS

### Study setting and period

The study was conducted in the Goba district, Bale zone, Oromia Regional State, Ethiopia. Goba town is located 446 km southeast of Addis Ababa (the capital city of Ethiopia) and 12 km southeast of Robe, the capital city of the Bale zone. The Goba district has one town administration and one rural woreda. According to the 2021 Goba district health office report[13], the total population of the Goba district is 115,857 (56,790 males and 59,067 females), and 3.47% (4,020) of the population are pregnant women. The district has 6 health centers (2 urban and 4 rural health centers). The district has 26 health posts and 6 primary clinics, 6 medium clinics, 9 drug stores, and 2 pharmacies [13].

### Study design and population

An institutional-based cross-sectional study design was used. The participants were pregnant women who attended antenatal care follow-up at public health facilities in the Goba district. The study participants were all randomly selected pregnant women who attended public health facilities found in Goba district during the study period and fulfilled the inclusion criteria. The study population consisted of a selected pregnant woman who lived for at least 6 months in the study area and who attended antenatal care follow-up at selected public health facilities during the study period. Pregnant women who lived for less than six months in the study area and those who were critically ill were excluded from the study. The study was conducted from May-June; 2022.

### Sample size determination and sampling technique

#### Sample size determination

The sample size was computed using a single population proportion for the primary objective by using Epi Info version 7.2.5.0 statistical tool and using the prevalence of undernutrition among pregnant women taken from a study conducted in Malga district, southern Ethiopia 26% [14], and by considering the following assumptions:

- CI =95%,
- Margin of error =5%
- P= Prevalence of undernutrition among pregnant women in Malga district, southern Ethiopia 26% [14].
- Using a design effect of 1.5, the calculated sample size was 443; by adding a 10% nonresponse rate, the final sample size was 487.

For the sample size required to address the second specific objective, the double population proportion formula was used to determine the factors associated with undernutrition among pregnant mothers exposed and not exposed to undernutrition based on a systematic review and meta-analysis performed in Africa [15], as displayed in(Supplemental Table 1).

n = (Z_a/2_+Z_β_) ^2^ × (p_1_ (1-p_1_) + p_2_ (1-p_2_))/(p_1_ –p_2_)^2^

#### Sampling technique and sampling procedure

A multistage stratified sampling technique was used, followed by a systematic random sampling technique to reach each study participant. Health centers found in Goba district were initially stratified into urban and rural (2 urban and 4 rural health centers); then, one health center from urban areas and two health centers from rural areas were selected by the lottery method from a total of 6 health centers (50% of the health centers). The selected health centers were Harawa Sinja Health Centre, which is located in an urban area; Misra; and Wecho Health Centre, which is located in a rural area.

The number of study subjects was allocated according to each health facility’s number of pregnant women who attended ANC in each health center every six months multiplied by the total sample size (n= 487) divided by the total number of pregnant women who attended the ANC unit of the selected public health facilities (N= 1,113). Finally, the study participants were selected from each health center using a systematic random sampling technique; the study participants were selected by the lottery method.

In general, the sampling technique was as follows: first, the most recent six months number of pregnant women who attended ANC follow-ups at the three selected health centers were gathered and divided by six to obtain the average one-month data for the purpose of sample allocation and sharing. The one-month average number of pregnant women data collected from the selected health centers using ANC registration logbooks was as follows: 218 from the Misra Health Centre, 229 from the Wecho Health Centre, and 666 from the Harawa Sinja Health Centre.

As a result, the following computation was used to determine the sample sizes of the health centers:

The total number of pregnant women obtained from all health centers is N= **1,113.**

✔ For the Misra health center: (218/1,113)*100% = 20%, sample sharing with the health center = (20/100) *487= **97**
✔ For the Wecho health center: (229/1,113)*100% = 20%, sample sharing with the health center = (20/100)*487 = **97**
✔ For the Harawa Sinja health center: (666/1,113) 100% = 60%, sample sharing to the health facility = (60/100)*487 = **293**

Study participants were chosen using a systematic random sampling procedure at every K^th^ (2^nd^) interval (K is the sample fraction; N/n = 1,113/487= 2), as shown in (Supplemental Figure 1).

#### Data collection procedures (tools, techniques and personnel)

The data were collected by using an interviewer-administered questionnaire. Age, marital status, residence, occupation, maternal educational status, religion, and husband educational status were collected, as well as respondents’ obstetric history, nutritional knowledge, dietary diversity, and anthropometric measurements. Sociodemographic variables were adapted from the Ethiopian demographic and health survey (EDHS 2016) [16]. The standard tool for measuring the wealth index was adopted from the EDHS (Ethiopian demographic and health survey 2016). Maternal anthropometric measurements were done according to the standards. MUAC of a pregnant mother was measured at mid-point between the tip of the shoulder and the tip of the elbow of the left arm. An adult Mid-Upper Arm Circumference (MUAC) tape that was non-elastic and non-stretchable was used to take measurements, after checking that the tape was applied with correct tension. The Mid-Upper Arm Circumference (MUAC) of a pregnant mother was read and documented to the nearest 0.1cm. MUAC measurement was performed by clinical health professionals following standard instructions and steps. A range below 23 cm was an indicator of Undernutrition and a range of >23 cm was for normal nutritional status.

A dietary diversity questionnaire adopted from FANTA 2006 was used to determine the dietary diversity status of the pregnant women. The number of food groups consumed before 24 hours before the survey was assessed. The foods consumed in the preceding 24-hour recall were grouped into 9 food groups. The dietary diversity score (DDS) was calculated from a single 24-hr recall method, and all the foods and liquids consumed a day before the study were categorized into 9 food groups. Consuming a food item from any of the groups was assigned a score of 1; if no food was consumed, a score of 0 was given. The nine food groups used to measure WDDSs were Starchy staples, Dark green leafy vegetables, Other fruits and vegetables, Organ meat, Meat and fish, Eggs, Legumes, nuts and seeds, and milk and milk products. The WDDS reflects the probability of micronutrient adequacy in the diet; therefore, the food groups included in the score are tailored for this purpose[17]. Six midwife professionals working in the ANC of Goba district health centers were recruited. The data collectors were responsible for the interviews and anthropometric measurements (Supplemental File 1).

### Study variables

#### Dependent variable

➢ **For the nutritional status of pregnant women**, a MUAC range less than 23 cm is an indicator of undernutrition, and a range greater than or equal to 23 cm (≥ 23 cm) is considered to indicate normal nutritional status [18].

#### Independent variable

**Socio demographic factors** included maternal age, marital status, maternal educational status, husband’s education, occupation, residence, family size, and wealth index).

**Dietary habits and dietary diversity-related factors (**nutritional knowledge, dietary diversity, MUAC of pregnant women, meal frequency, skipping meals to avoid weight gain, and iron-foliate supplementation)

**Economic factor (**wealth index)

### Operational definition

#### Undernutrition

Pregnant women with a MUAC< 23 cm [18].

#### High dietary diversity score

Pregnant women who consumed 7-9 food groups in the 9-food group were categorized as having a high dietary diversity score [17].

#### Medium dietary diversity score

Pregnant women who consumed 4-6 food groups out of the 9 food groups were categorized as having a low dietary diversity score [17].

#### Low dietary diversity score

Pregnant women who consumed < 3 food groups out of the 9 food groups were categorized as having a low dietary diversity score[17].

#### Good dietary knowledge

Pregnant women scored with the highest tertile score (≥8) of the 12 nutrition-related knowledge questions[19].

#### Poor dietary knowledge

Pregnant women scored with the lowest two tertile scores (<8) on the 12 nutrition-related knowledge questions[19].

### Data quality assurance

Data quality control measures were undertaken to increase the reliability of the outcome. The questionnaire was prepared in English and translated into Afan Oromo and back-translated to English by language experts to check its consistency. The data collectors and supervisors were given two days of training. Initially, the investigator checked for appropriateness, such as typing errors, missing questions, and inappropriateness. Pretesting was performed on pregnant women attending ANC at one of the health centers in the Bale zone, corresponding to 5% of the calculated sample size, to check the validity and reliability of the data collection tools, and necessary modifications were made accordingly. The data collectors submitted the collected data on a daily basis to the supervisors and the investigator. Each questionnaire was checked before data entry for completeness and consistency. Two different data collectors took MUAC measurements, and average measurements were taken for analysis.

Regarding the dietary diversity of pregnant women, lists of locally available food were adapted by interviews with key informants and agriculture research experts in the nearby study area. The pregnant mother was asked to recall the foods they had consumed in the previous 24 hours, followed by probing to ascertain that no meal or snack was left out.

### Data processing and analysis

The data were checked for completeness and consistency, and Epi-Data v4.6.0.4 was used to perform the cleaning. The cleaned data were exported to SPSS version 24 for statistical analysis.

First, descriptive statistics were computed, and the results are reported as frequencies and percentages, tables, and graphs. Next, bivariate logistic regression was performed, and variables with a p value < 0.25 were subjected to multivariable logistic regression to identify the factors associated with nutritional status among pregnant mothers. Finally, variables with a P value < 0.05 in the multivariable logistic regression model were considered to be statistically significant, and an adjusted odds ratio with its 95% confidence interval was used to determine the association. Multicollinearity among the independent variables was tested using the variance inflation factor (VIF).

The model fitness of the test was checked by the Hosmer–Lemeshow goodness of fit test. Categorical variables such as dietary knowledge (good/poor) and the nutritional status of the pregnant women (undernutrition/normal) are summarized as the frequency and percentage in the table.

For the MUAC of pregnant women, a range below 23 cm is an indicator of undernutrition, and a range ≥ 23 cm is considered to indicate normal nutritional status. A DDS of 9 points was computed for the pregnant mothers by adding the values of all the groups. The pregnant women were then categorized as low (≤ 3), medium (4-6), or high (7-9). Principal component analyses (PCAs) were performed for the household wealth score, and the household wealth index was ranked into tertiles.

### Ethical considerations

The study protocol was approved, and the Ethical Review Committee of Salale University provided ethical approval letter with reference number HSC/878/14. The study was performed in accordance with the World Medical Association Declaration of Helsinki on medical research. Informed verbal consent was obtained from every study subject before the data collection. The participant’s right to discontinue or leave the study was also secured. The entire information collected from the study participants was handled confidentially by omitting their identification.

### Patient and public involvement

Patients or the public were not involved in the study design, conduct, reporting or distribution strategies of the research.

## RESULTS

### Socio demographic and economic characteristics of the study participants

In total, four hundred eighty-seven pregnant women were recruited for this study, and the response rate was 100%. The age of the study participants ranged from 15-45 years. Of these, 144 (29.6%) were aged between 20 and 24 years, and 61 (12.5%) were aged 15 and 19 years. The mean (±SD) age of the study participants was 27.01 (±6.59) years. Regarding residency, 293 (60.2%) of the respondents were living in urban areas.

The dominant ethnic group of the study participants was Oromo 365 (74.9%), followed by Amhara 105 (21.6%). Regarding religion, 232 (47.6%) was Orthodox. Majority of the study participants were married (96.7%), as shown in (Table 1).

**Table 1:**
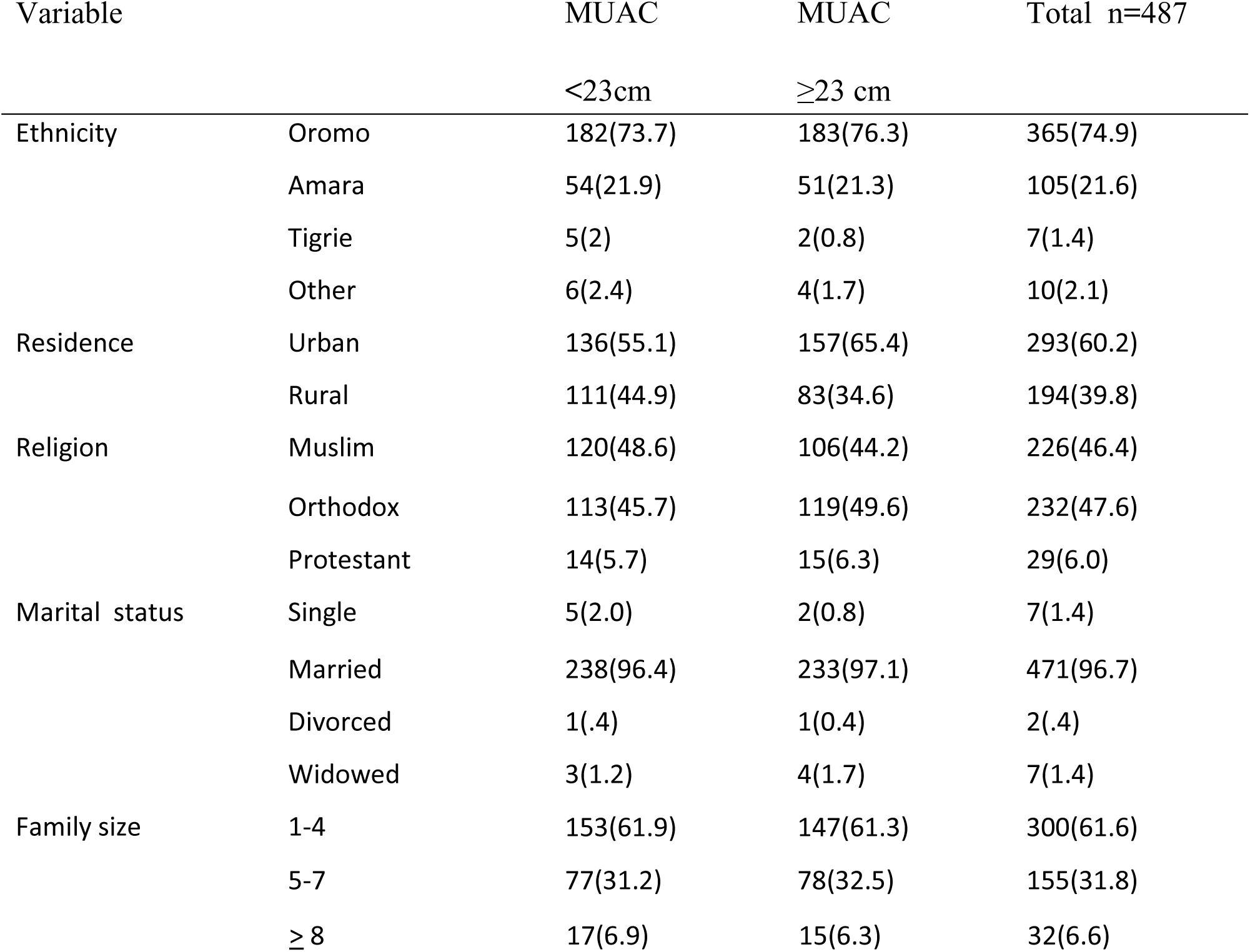
Socio-demographic characteristics of pregnant mothers in Goba District from May-June, 2022, (N = 487)

Regarding maternal educational status, 280 (57.5%) had no formal education, and 69 (14.2%) had a secondary education, as displayed in (Supplemental Figure 2).

### Nutrition-related factors

The majority of the study participants (66.7%) used Teff Injera and wet (Ethiopian stew) as staple food. A total of 227 (46.6%) of the respondents had a meal frequency of 4-5 times per day. Regarding food taboos, 15 (3.1%) of the study participants had experienced food restriction during pregnancy due to cultural beliefs. Regarding tea/coffee consumption, 450 (92.4%) participants were taking tea/coffee immediately after eating food every day. Concerning appetite status during pregnancy, 178 (36.6) participants experienced appetite loss during pregnancy, as displayed in (Table 2).

**Table 2:**
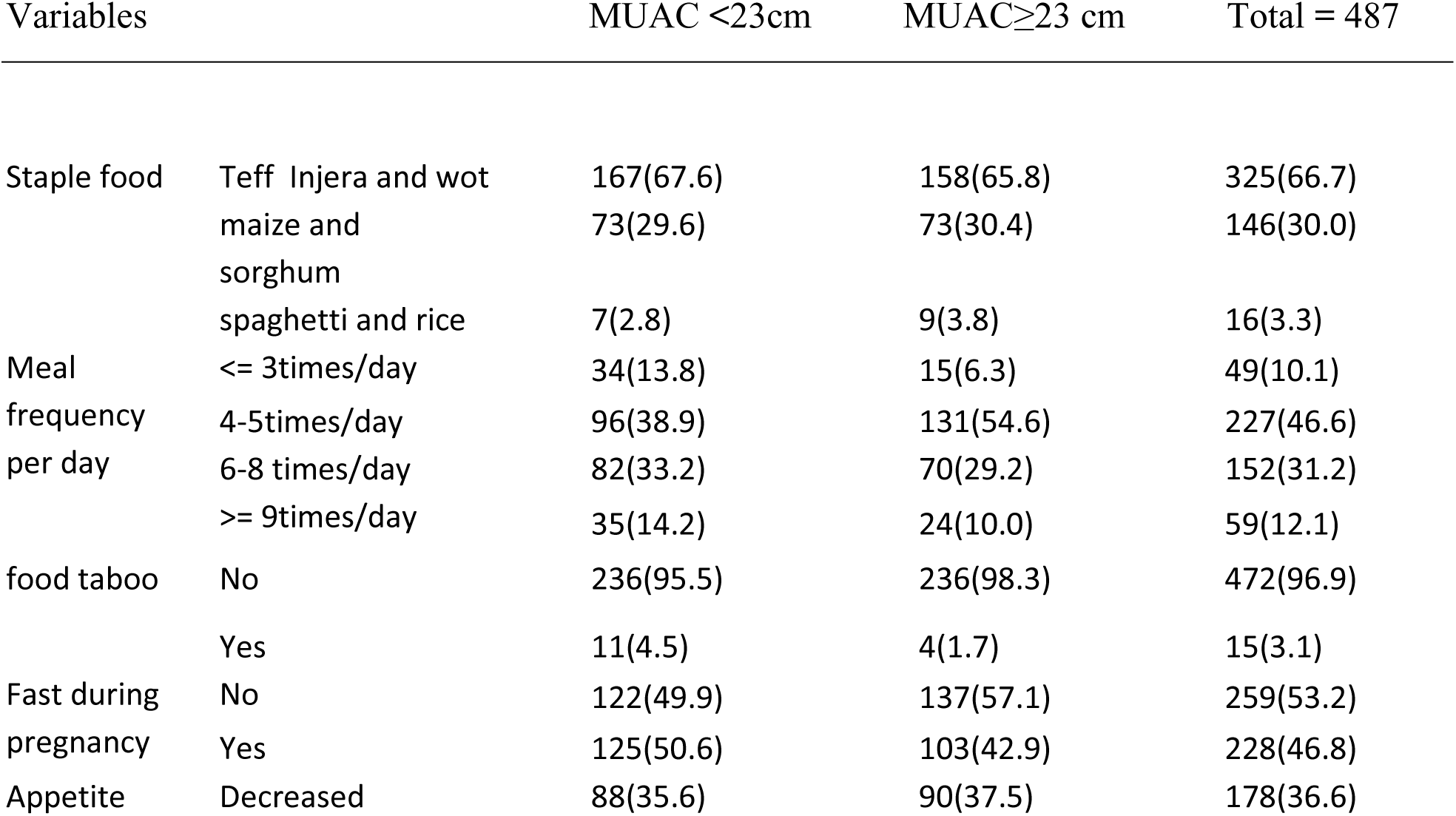
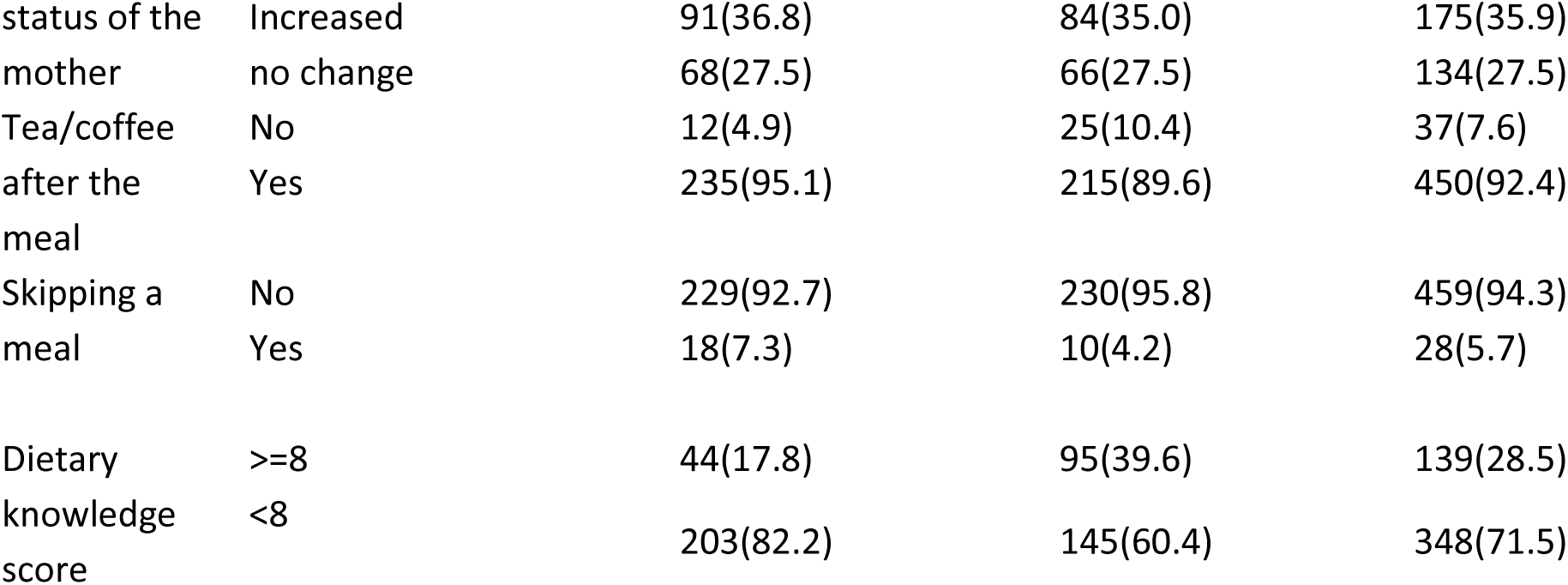
Dietary diversity, Dietary habits, and Nutrition related factors among pregnant mothers in Goba district selected public health facilities from May-June, 2022, (N = 487).

### Magnitude of Undernutrition

The magnitude of undernutrition among pregnant women in public health facilities of the Goba district was 50.7%. The mean MUAC (±SD) of the study participant was 24.55(±3.59). The MUAC range of Study participants was 20 cm - 37 cm, as shown in (Figure 1).

### Dietary diversity score

Regarding maternal dietary diversity scores, 181 (37.2%) pregnant mothers had a low dietary diversity score (≤ 3 food groups), and only 29 (6.0%) pregnant mothers had a high dietary diversity score, as displayed in (Figure 2).

### Maternal health and illness-related factors

Regarding maternal health and illness-related factors, 27.3% of the study participants had a previous history of abortion, and 9% of them had a history of stillbirth. Of the total study participants, 162 (33.3%) had >= 4 pregnancies. The majorities (53.8%) of the respondents were in the first trimester of pregnancy, and only 23.4% of them were supplemented with iron folic acid tablets, as displayed in (Table 3).

**Table 3:** Maternal health and illness-related factors among pregnant mothers in Goba district selected public health facilities from May – June, 2022, (N= 487)

| Variables |  | MUAC | MUAC | Total n=487 |
| --- | --- | --- | --- | --- |
|  |  | <23 cm | ≥23 cm |  |
| Anemia in previous pregnancy | No | 205(83.0) | 192(80.0) | 397(81.5) |
|  | Yes | 42(17.0) | 48(20.0) | 90(18.5) |
| Iron-folic supplementation | No | 53(21.5) | 61(25.4) | 114(23.4) |
|  | Yes | 194(78.5) | 179(74.6) | 373(76.6) |
| felt illness in your current pregnancy | No | 151(61.1) | 195(81.3) | 346(71.0) |
|  | Yes | 96(38.9) | 45(18.8) | 141(29.0) |
| history of Stillbirth | No | 222(89.9) | 221(92.1) | 443(91.0) |
|  | Yes | 25(10.1) | 19(7.9) | 44(9.0) |
| Gestational age of pregnancy | 1st Trimester | 135(54.7) | 127(52.9) | 262(53.8) |
|  | 2nd Trimester | 68(27.5) | 67(27.9) | 135(27.7) |
|  | 3rd Trimester | 44(17.8) | 46(19.2) | 90(18.5) |
| No. of pregnancy category | <= 3 | 174(70.4) | 151(62.9) | 325(66.7) |
|  | >= 4 | 73(29.6) | 89(37.1) | 162(33.3) |
| History of Abortion | No | 169(68.4) | 185(77.1) | 354(72.7) |
|  | Yes | 78(31.6) | 55(22.9) | 133(27.3) |

### Factors associated with undernutrition

Finally, six variables (educational status of the mother, dietary knowledge status, number of meals the mother had per day, history of illness, and household wealth status) with p value < 0.05 were the independent predictors of maternal undernutrition. An additional file shows this in more detail (Supplemental Table 2).

The present study showed a significant association between pregnant mothers’ undernutrition and dietary knowledge. The risk of undernutrition for pregnant women who had poor dietary knowledge (AOR= 5.588, 95% CI (2.921-10.689)) was 5.588 higher than that for pregnant women who had good dietary knowledge. A significant association was also observed between undernutrition and the frequency of eating a meal per day. The risk of undernutrition in pregnant mothers who had a meal frequency <= 3 (AOR = 3.292; 95% CI (1.040-10.422) was three times greater than that in pregnant women who had a meal frequency >3 times per day. Another significant factor associated with undernutrition in pregnant mothers was having a history of illness during pregnancy. A pregnant mother who had a previous history of illness/disease during pregnancy (AOR= 2.089, 95% CI (1.246-3.504)) was 2.089 times more likely to be undernourished than was a pregnant mother who had no history of illness (Table 4).

**Table 4:** Multivariable logistic regression analysis of factors associated with undernutrition among pregnant mothers in Goba district from May-June, 2022, (N = 487)

| predictors | | MUAC<br>< 23cm | MUAC<br>$\geq 23$cm | COR | AOR | 95 % C.I | | p-value |
| --- | --- | --- | --- | --- | --- | --- | --- | --- |
|  |  |  |  |  |  | lower | upper |  |
| Educational status of mother | No formal education | 12(4.9) | 80(33.3) | .057 | 5.050 | 1.470 | 17.346 | .010** |
|  | Grade 8 & Below | 16(6.5) | 53(22.1) | .115 | .220 | .056 | .863 | .030** |
|  | High school complete | 11(4.5) | 18(7.5) | .232 | .477 | .122 | 1.859 | .286 |
|  | College diploma | 5(2.0) | 12(5.0) | .158 | 1.125 | .261 | 4.856 | .874 |
|  | Degree and above | 12(4.9) | 80(33.3) | 1 |  |  |  |  |
| Dietary knowledge | Good | 44(17.8) | 95(39.6) | 1 |  |  |  |  |
|  | poor | 203(82.2) | 145(60.4) | 3.023 | 5.588 | 2.921 | 10.689 | .000 |
| Meal frequency | $\leq 3$times/day | 34(13.8) | 15(6.3) | 1.554 | 3.292 | 1.040 | 10.422 | .043** |
|  | 4-5times/day | 96(38.9) | 131(54.6) | .503 | 1.201 | .452 | 3.189 | .713 |
|  | 6-8 times/day | 82(33.2) | 70(29.2) | .803 | .566 | .226 | 1.419 | .225 |
| | $\geq 9$times/day | 35(14.2) | 24(10.0) | 1 | | | | |
| History of illness | No | 151(61.1) | 195(81.3) | 1 |  |  |  |  |
|  | Yes | 96(38.9) | 45(18.8) | 2.755 | 2.089 | 1.246 | 3.504 | .005 |
| No of pregnancy | $\leq 3$ | 174(70.4) | 151(62.9) | 1 | | | | |
| | $\geq 4$ | 73(29.6) | 89(37.1) | 1.405 | .582 | .342 | .989 | .045 |

| category |  |  |  |  |  |  |  |  |
| --- | --- | --- | --- | --- | --- | --- | --- | --- |
| HH wealth | Poor | 101(40.9) | 80(33.3) | 2.806 | 4.774 | 2.216 | 10.285 | .000** |
|  | Medium | 137(55.5) | 140(58.3) | 2.175 | 2.219 | 1.133 | 4.346 | .020 |
|  | Rich | 32(13.0) | 67(27.9) | 1 |  |  |  |  |

## Discussion

This study assessed the magnitude and associated factors of undernutrition among pregnant women attending ANC services at public health facilities in the Goba district. The present study identified the importance of undernutrition for public health in the study area.

More than half (50.7%) of the pregnant women were undernourished. Almost similar findings were reported from a study conducted in Kacha Birra district, southern Ethiopia (52.6%)[20]. A poor nutritional status was reported for 50% of the pregnant women from Dhaka, Bangladesh[21]. The similarity might be attributed to the socioeconomic status of the study participants.

However, these findings were greater than those of a study conducted at Alamata General Hospital, Northern Ethiopia, which was reported to be 23.2%[22]. A study in the Silte zone, reported 21.8%[23]. A meta-analysis and systematic review performed in Ethiopia, which identified a pooled prevalence of 29.07% malnutrition among pregnant women in Africa[15]. A possible explanation for this discrepancy might be socioeconomic and cultural variation.

Conversely, a study conducted in Aleta Chuko, southern Ethiopia, reported that the prevalence of undernutrion among pregnant mothers was 71.15%. This might be because of the difference in the Sociodemographic and economic status of the study participants.

Mothers who had no formal education (who could not read or write) were 5 times more likely to become undernourished than mothers whose educational status was a degree or above. A study from Mettu, Southwest Ethiopia, reported greater odds of undernutrition among pregnant women without a formal education[24]. This is certainly because the more mothers have sufficient education, the better their maternal nutritional status is. In contrast, a study performed in Alemata, Northern Ethiopia, reported a lower risk of undernutrition among pregnant women without formal education than among those with a university degree[22]. This discrepancy might be due to variations in Sociodemographic and economic conditions.

In the present study, mothers’ poor dietary knowledge was significantly associated with undernutrition. The odds of undernutrition were 6 times greater for mothers who had poor dietary knowledge than for mothers who had good dietary knowledge. This finding is in agreement with a study performed in Dessie town, Northern Ethiopia, in which higher odds of maternal undernutrition were reported among pregnant women with poor nutritional knowledge[25]. This finding was also supported by the findings from the Silte zone, southern Ethiopia[23]. Similar finding was reported by the findings from Bench-Sheko and Kaffa zones; southwestern Ethiopia[26]. A possible explanation for this might be that good knowledge about basic nutrients and diversified diets results in good dietary practices, which are important for optimum health, especially during pregnancy.

The study further showed that meal frequency was significantly associated with maternal undernutrition. Pregnant mothers who had a meal frequency of less than or equal to three times per day were more likely to be undernourished. These findings are consistent with the findings of a study performed in Illu Ababor, Southwest Ethiopia, which reported greater odds of undernutrition among pregnant women consuming fewer than three meals per day[19]. This is because having eaten less than or equal to three times a day during pregnancy results in a lack of diversified food and low-energy food for pregnant mothers, which might ultimately result in undernutrition[27].

The odds of undernutrition were 2.089 times greater among pregnant mothers who had a history of illness during pregnancy than among pregnant women who had no history of illness during pregnancy. This finding is consistent with the findings of Dissie town, Northern Ethiopia, which reported that the prevalence of undernutrition was 93% greater in women who had a history of illness during pregnancy [25]. This is because illness results in increased energy and nutrient demand and decreased appetite (loss of appetite), which ultimately results in undernutrition.

However, a study performed in the Bench-Sheko and Kaffa zones, Southwest Ethiopia, reported that any illness during pregnancy was not associated with pregnant mothers’ undernutrition [26]. The variation might be due to differences in the pre pregnancy and post pregnancy health-seeking behaviors of the study participants.

Pregnant women from households with lower wealth status were 4.774 times more likely to develop undernutrition than were those from households with the highest wealth status. The finding is in agreement with those of a study done in the Illu Aba Bor zone, Southwest Ethiopia, which reported greater odds of maternal undernutrition among women from the lowest wealth quintile [19]. This result is also supported by a study of 23 African countries that indicated a lower risk of maternal malnutrition among those with a better household economy [15]. A possible reason for this might be that women with higher economic status have an increased chance of purchasing power and can afford to eat a diversified diet, which in turn improves their nutritional status.

This study was inconsistent with the study performed at the University of Gondar Hospital, Southwest Ethiopia, which reported that household wealth status was not associated with undernutrition among pregnant mothers [28]. This discrepancy might be due to the study setting (only one health facility) and the socioeconomic status of the study population.

In the present study, a significant association was observed between parity and undernutrition. The prevalence of undernutrition was significantly greater among pregnant women who had more than or equal to four pregnancies than among pregnant mothers who had less than or equal to three. These findings are consistent with the findings of studies performed at the University of Gonder Hospital, Northwest Ethiopia, which reported a greater proportion of undernutrition among pregnant women who had >= 4 pregnancies [28]. These findings were also supported by a meta-analysis and a systematic review conducted in Ethiopia, which reported that as parity increases, maternal malnutrition also increases[29]. A greater number of children in the household obligate the mother to take care of her children rather than taking care of her health, and a greater number of children results in limited household resources. However, there are a number of limitations which should be considered. First, social desirability bias and recall bias could affect the accuracy of the data, particularly for the 24-hour dietary diversity recall and illness history. Second, the study excluded pregnant women who were not utilizing ANC services. Lastly, this study is cross-sectional and was analyzed descriptively, meaning that no causal inferences can be drawn.

## Conclusion

This study revealed that the magnitude of undernutrition among study participants in the study area was high. The educational status of the mother, poor dietary knowledge, number of meals a mother had per day, history of illness during pregnancy, and household wealth status were found to be significantly associated with undernutrition among pregnant mothers attending ANC services at public health facilities. Therefore, further community-based studies with larger sample sizes should be conducted to identify other risk factors and associated factors of undernutrition among pregnant women to address those women who are unable to attend antenatal care clinics in health facilities. Undernutrition problems should be addressed through the implementation of integrated preventive strategies. For example, with respect to women’s education, nutritional care should be integrated into maternity services (behavioral change communication should be enhanced for pregnant women, dual counseling services for pregnant women and her husband should be provided during ANC services, and sustained nutritional education to enhance the nutritional knowledge and practices of pregnant women should also be sought. This study recommends that the Woreda and town health offices plan and implement nutrition education and promotion programs for women of reproductive age and pregnant women. Pregnant women should be encouraged to eat more diverse foods and increase their meal and snack intake. Healthcare providers should provide thorough nutrition education and counseling to pregnant women, strengthen dual counseling with husbands during antenatal care, ensure proper follow-up and treatment of diseases during pregnancy, and promote family planning. Finally, future research should conduct larger community-based studies to identify additional risk factors for undernutrition among pregnant women.

## Ethical approval and consent to participate

Ethical approval was received from the Ethical Review Committee of Salale University with reference number HSC/878/14. The study was performed in accordance with the World Medical Association Declaration of Helsinki on medical research. It was presented to the Goba district health office and selected health facilities to obtain official permission to undertake research activities at each of the selected health facilities. All study participants were informed that the data were kept private and confidential and used only for research purposes. The participants were assured that they had the right to refuse or withdraw if they were not comfortable at any time. Informed verbal consent was obtained from all participants and from the parent and/or legal guardian for participants younger than 16 years of age before the interview.

## Consent for publication

Not applicable

## Data availability statement

The original contributions presented in the study are included in the article/supplementary material, and further inquiries can be directed to the corresponding author/s.

## Competing interests

The authors declare that they have no competing interests.

## Funding

The authors received no funding for this research.

## Author contributions

Conceptualization: S.M.I. and M.S.L. Data curation: S.M.I. and M.S.L. Formal analysis: S.M.I., M.S.L., D.H., H.K., and H.A. Investigation: Project administration: M.S.I., S.M.L., A.F.A., D.H. and H.A. Software: M.S.L., A.F.A, S.M.I, H.K., H.A. and D.H. Validation: M.S.L., A.F.A., S.M.I. and D.H. Visualization: S.M.I. and M.S.L. Writing and original draft of manuscript: S.M.I. and M.S.L. Writing, review and editing of manuscript: All authors. S.M.I. is responsible for the overall content as the guarantor.

## Acknowledgment

First and foremost, we would like to thank all the study participants for their participation in this study. Second, we would like to acknowledge Salale University, Department of Public Health, for the approval of the ethical clearance for this study. Finally, we also thank the Bale Zone Health Office and the Goba District Health Office for providing baseline information.

## Abbreviations

ANC: Ante Natal Care
AOR: Adjusted Odds Ratio
BMI: Body Mass Index
CI: Confidence Interval
COR: Crude Odds Ratio
EDHS: Ethiopian Demographic Health Survey
EMDHS: Ethiopian Mini Demographic Health Survey
FAO: Food and Agricultural Organization
LBW: Low Birth Weight
MUAC: Mid-Upper Arm Circumference
SPSS: Statistical Package for Social Sciences
WDDS: Women’s Dietary Diversity Score

## Author details

Department of Public Health, Ginnir General Hospital, Oromia Regional Health Bureau, Ginnir, Ethiopia

Department of Public Health, Goba District Health Office, Oromia Regional Health Bureau, Goba, Ethiopia

Department of Public Health, College of Health Science, Salale University, Fitche, Ethiopia

Department of Nursing, Kuyu General Hospital, Oromia Regional Health Bureau, Gerba Guracha, Ethiopia

Department of Public Health, International Rescue Committee Adama Field Office, Adama, Ethiopia

## Figure legends

**Figure 1:** Magnitude of Undernutrition among pregnant mothers attended ANC at public health facilities of Goba district, Bale zone, Southeast Ethiopia, 2022. The red part indicates pregnant women who were undernourished or MUAC < 23 cm, and the blue part indicate pregnant women who were in normal range for nutritional status or their MUAC >= 23cm.

**Figure 2:** Dietary diversity score of pregnant mothers attending ANC at public health facilities of Goba District South east Ethiopia, 2022. The light Green shaded part of the Pie chart (6.0%) indicates pregnant women with high dietary diversity score: Pregnant women who consumed 7-9 food groups in the 9-food group were categorized as having a high dietary diversity score. The Red shaded part of the Pie chart (56.9%) indicates pregnant women with medium dietary diversity score: Pregnant women who consumed 4-6 food groups out of the 9 food groups were categorized as having a low dietary diversity score.

The Blue shaded part of the Pie chart (37.2%) indicates pregnant women with Low dietary diversity score: Pregnant women who consumed ≤ 3 food groups out of the 9 food groups were categorized as having a low dietary diversity score.

## Notes

### Competing Interest Statement

The authors have declared no competing interest.

### Author Declarations

The study protocol was approved, and the Ethical Review Committee of Salale University provided ethical approval letter with reference number HSC/878/14.

