## Supplemental Table 1 for "Magnitude of Undernutrition and Associated Factors among Pregnant Women Attending Public Health Facilities in Goba District, Bale Zone, Oromia, Ethiopia: A Cross-sectional Study"

Supplemental Table 1: Sample size calculation based on the double population proportion formula

| Variables | Assumptions | | | | | Sample size |
|---|---|---|---|---|---|---|
| | Proportion | | Power | Confidence interval | Ratio | |
| | P1 | P2 | | | | |
| Rural residency | 25.8% | 51.3% | 80% | 95% | 1:1 | 160 |
| Low husbands education | 23.5% | 52.5% | 80% | 95% | 1:1 | 100 |
| Multiple pregnancy | 20.6% | 45.5% | 80% | 95% | 1:1 | 126 |
| Marital status | 60% | 40% | 80% | 95% | 1:1 | 214 |
