## Supplemental Table 2 for "Magnitude of Undernutrition and Associated Factors among Pregnant Women Attending Public Health Facilities in Goba District, Bale Zone, Oromia, Ethiopia: A Cross-sectional Study"

Supplemental Table 2: Bivariate analysis of study participants among pregnant women in Goba district selected public health facilities from May – June, 2022, (N = 487)

| variables | | MUAC < 23 cm | MUAC ≥23 cm | COR | 95% C.I lower | upper | P-value |
|---|---|---|---|---|---|---|---|
| Age | 15-19 | 44(17.8) | 17(7.1) | 1.941 | .946 | 3.984 | .071* |
| | 20-24 | 68(27.5) | 76(31.7) | .671 | .384 | 1.172 | .161 |
| | 25-29 | 60(24.3) | 72(30.0) | .625 | .355 | 1.101 | .104 |
| | 30-34 | 31(12.6) | 42(17.5) | .073 | .554 | .290 | .073 |
| | ≥35 | 44(17.8) | 33(13.8) | 1.333 | - | - | .212 |
| residence | Urban | 136(55.1) | 157(65.4) | 1.337 | - | - | .045 |
| | Rural | 111(44.9) | 83(34.6) | .648 | .449 | .934 | .020* |
| marital status | Single | 5(2.0) | 2(0.8) | 1 | | | |
| | Married | 238(96.4) | 233(97.1) | 3.333 | .362 | 30.701 | .288 |
| | Divorced | 1(.4) | 1(0.4) | 1.362 | .302 | 6.152 | .688 |
| | Widowed | 3(1.2) | 4(1.7) | 1.333 | .057 | 31.121 | .858 |
| Family size | 1-4 | 153(61.9) | 147(61.3) | 1 | | | .724 |
| | 5-7 | 77(31.2) | 78(32.5) | .918 | .442 | 1.906 | .819 |
| | ≥8 | 17(6.9) | 15(6.3) | .871 | .406 | 1.867 | .723 |
| Meal frequency | <= 3times/day | 34(13.8) | 15(6.3) | 1.554 | .699 | 3.457 | .280 |
| | 4-5times/day | 96(38.9) | 131(54.6) | .503 | .281 | .900 | .021* |
| | 6-8 times/day | 82(33.2) | 70(29.2) | .803 | .437 | 1.478 | .481 |
| | >= 9times/day | 35(14.2) | 24(10.0) | 1 | | | |
| Staple food of mother | Teff Injera and wot | 167(67.6) | 158(65.8) | | | | |
| | maize and sorghum | 73(29.6) | 73(30.4) | 1.359 | .494 | 3.736 | .552 |
| | spaghetti and rice | 7(2.8) | 9(3.8) | 1.286 | .455 | 3.636 | .636 |
| food taboo | No | 236(95.5) | 236(98.3) | 1 | | | |
| | Yes | 11(4.5) | 4(1.7) | 2.750 | .863 | 8.759 | .087* |
| Tea/coffee | No | 12(4.9) | 25(10.4) | 1 | | | .346 |
| | Yes | 235(95.1) | 215(89.6) | .439 | .215 | .896 | .024* |
| History of abortion | No | 169(68.4) | 185(77.1) | | | | |
| | Yes | 78(31.6) | 55(22.9) | 1.552 | 1.037 | 2.323 | .033* |

| | | | | | | | |
|---|---|---|---|---|---|---|---|
| History of illness | No | 151(61.1) | 195(81.3) | 1 | | | |
| | Yes | 96(38.9) | 45(18.8) | 2.755 | 1.823 | 4.163 | .000* |
| Fast during pregnancy | No | 122(49.9) | 137(57.1) | 1 | | | |
| | Yes | 125(50.6) | 103(42.9) | 1.363 | .954 | 1.948 | .089* |
| Appetite status of mother | Decreased | 88(35.6) | 90(37.5) | 1.108 | .730 | 1.682 | .630 |
| | Increased | 91(36.8) | 84(35.0) | 1 | | | |
| | no change | 68(27.5) | 66(27.5) | 1.054 | .673 | 1.650 | .819 |
| Gestational age | 1st trimester | 135(54.7) | 127(52.9) | 1 | | | |
| | 2nd trimester | 68(27.5) | 67(27.9) | 1.111 | .688 | 1.794 | .666 |
| | 3rd trimester | 44(17.8) | 46(19.2) | 1.061 | .622 | 1.809 | .828 |
| No of pregnancy category | < =3 | 174(70.4) | 151(62.9) | | | | |
| | ≥ 4 | 73(29.6) | 89(37.1) | 1.405 | .962 | 2.051 | .078* |
| DDS | Low | 101(40.9) | 80(33.3) | 2.806 | 1.212 | 6.496 | .016* |
| | Medium | 137(55.5) | 140(58.3) | 2.175 | .957 | 4.943 | .064* |
| | High | 9(3.6) | 20(8.3) | 1 | | | |
| HH wealth status | Poor | 110(44.5) | 66(27.5) | 3.490 | 2.074 | 5.870 | .000* |
| | Medium | 105(42.5) | 107(44.6) | 2.055 | 1.246 | 3.387 | .005* |
| | Rich | 32(13.0) | 67(27.9) | 1 | | | |
| Anemia in previous pregnancy | No | 205(83.0) | 192(80.0) | 1 | | | |
| | Yes | 42(17.0) | 48(20.0) | 1.220 | .771 | 1.930 | .395 |
| Iron-folic supplementation | No | 53(21.5) | 61(25.4) | 1 | | | |
| | Yes | 194(78.5) | 179(74.6) | 1.247 | .819 | 1.899 | .303 |
| Skipped meal to avoid weight gain | No | 229(92.7) | 230(95.8) | | | | |
| | Yes | 18(7.3) | 10(4.2) | .553 | .250 | 1.224 | .144* |
| Dietary knowledge category | Good | 44(17.8) | 95(39.6) | 1 | | | |
| | Poor | 203(82.2) | 145(60.4) | 3.023 | 1.994 | 4.583 | .000* |

* Candidate variables for multivariable logistic regression
